# Reduced Frontal Cortical Tracking of Conflict between Selfish versus Prosocial Motives in Narcissistic Personality Disorder

**DOI:** 10.1101/2021.03.01.21252656

**Authors:** David S. Stolz, Aline Vater, Björn H. Schott, Stefan Roepke, Frieder M. Paulus, Sören Krach

## Abstract

Narcissistic Personality Disorder (NPD) entails severe impairments in interpersonal functioning that are likely driven by selfish and exploitative behavior. Here, we investigate the underlying motivational and neural mechanisms of prosocial decision-making by experimentally manipulating motivational conflict between selfish and prosocial incentives. One group of patients diagnosed with NPD and a group of healthy controls (CTL) were scanned using functional magnetic resonance imaging while performing a prosocial decision-making task. In this task, we systematically varied the level of conflict between selfish and prosocial options on each trial. We analyzed choice behavior, response times, and neural activity in regions associated with conflict monitoring to test how motivational conflict drives prosocial choice behavior. Participants in the NPD group behaved less prosocially than the CTL group overall. Varying degrees of motivational conflict between selfish and prosocial options induced response variability in both groups, but more so in the CTL group. The NPD group responded faster than the CTL group, unless choosing prosocially, which slowed response times to a level comparable to the CTL group. Additionally, neural activity tracking motivational conflict in dorsomedial prefrontal cortex was reduced in the NPD group. Collectively, low generosity in NPD appears to arise from reduced consideration of prosocial motives, which obviates motivational conflict with selfish motives and entails reduced activity in neural conflict monitoring systems. Yet, our data also indicate that NPD is not marked by an absolute indifference to others’ needs. This points to potentials for improving interpersonal relationships, effectively supporting the well-being of patients and their peers.

## Introduction

Narcissistic personality disorder (NPD) is characterized by a pervasive pattern of grandiosity, need for admiration, and lack of empathy[1]. Relationships in patients with NPD are described as largely superficial and serving as a means for self-esteem regulation, rather than being motivated by mutuality and genuine interest in the other person[1]. Most of these descriptive interpersonal features derive from clinical observations. Laboratory studies that prove these concepts using experimental designs are lacking[2]. The aim of this study was to investigate selfish behavior, a key interpersonal pattern in NPD, on a behavioral and neural level.

“Narcissism” can be conceptualized as a dimensional personality trait including grandiose and vulnerable features[3,4]. Grandiose features are also reflected in the descriptive DSM-5 criteria set of NPD[1] and include an exaggerated sense of self-importance and need for admiration. Vulnerable features include traits such as anger, shame, and insecurity[3]. A pervasive motif of grandiose narcissism is to maintain an exaggerated self-esteem, which is reflected in social behavior[5,6]. Non-clinically grandiose individuals overestimate their abilities[7], tend to be extraverted[8], and display interpersonal skills like charmingness[9,10], which presumably facilitates making new acquaintances[6,11,12]. In the long run, however, non-clinical grandiosity predicts more detrimental outcomes[5,12] reflected in antagonistic and defensive behavior[9,13,14], vindictiveness[15,16], and dysfunctional relationships[17,18]. Non-clinical narcissistic vulnerability similarly relates to defensiveness and self-enhancement[10], but also to emotional instability, low extraversion[19], low self-esteem[20], and aggression[21]. Most behavioral studies on “narcissism” are performed in non-clinical samples using the Narcissism Personality Inventory (NPI) that mainly captures grandiose features of the construct[21], making the transferability of these data into clinical samples disputable[22]. Pathological narcissism likely represents a singular dimension, with NPD as an extreme[23] that is linked to reduced empathy[24,25], and distress of close others[26].

In healthy subjects, laboratory studies have shown that higher trait narcissism predicts higher individual acquisitiveness but diminishes the group’s total harvest in common goods games[27]. Similarly, trait vulnerability predicts reduced generosity, lower sensitivity to potential punishment for low generosity, and increased retaliation[28]. This coheres with classic findings that non-clinical grandiosity predicted aggression, especially following self-esteem threats[29]. Yet, when prosocial motives gain salience through empathizing or perspective taking, both selfishness[30–32] and aggression[31,33] diminish in healthy individuals, and the arising arbitration between selfish and prosocial motives entails slower reaction times (RTs)[34–37]. Habitually selfish/prosocial individuals respond quickly when behaving selfishly/prosocially, whereas deviations from default behavior are slower[36]. Hence, to the extent that individuals inform their behavior by reconciling opposing interpersonal motives, experimentally induced conflict between such motives should slow down responses and potentially shift them away from default tendencies[34,36].

Regarding NPD, this evidence from non-clincal samples stimulates divergent hypotheses: Either, pronounced selfish behavior, as also frequently observed in non-clinical narcissism[27–29], could go along with low motivational conflict if selfish motives trump prosocial motives. Alternatively, if interpersonal deliberation in NPD entails motives like being perceived as likeable or attractive[9,10], achievable for instance through generous actions, eventually selfish behavior should demand to first resolve conflict with co-existing prosocial motives. In the first hypothesis, low motivational conflict should be reflected in fast RTs, whereas the second hypothesis would predict slower RTs[34,36].

Neurally, motivational conflict should relate to activity in areas of medial prefrontal cortex, such as dorsomedial prefrontal cortex (dmPFC), anterior cingulate cortex (ACC), and the supplementary motor complex (SMC, including supplementary motor area (SMA), and preSMA)[38]. These regions are commonly associated with cognitive control[39,40], which also holds in the social domain[41–43]. Further, since ventromedial prefrontal cortex (vmPFC) is associated with subjective value computations of competing choice options[44] and action outcomes[45], activity in this region may also contribute to reduced prosocial decision making in NPD. Moreover, the vmPFC tracks state self-esteem[46,47], social inequality assessment[48,49], and guides prosocial behavior in concert with the temporoparietal junction (TPJ)[37,50,51]. The TPJ includes regions at the border of temporal and parietal lobes including superior temporal sulcus (STS) and angular gyrus (AG)[52], implicated in higher-level social-cognitive processes[53,54] such as moral judgement[55] or perspective taking[30]. Last, the anterior insula (AI) is involved in interoceptive awareness[56], empathic processes[30,57], detection of social norm violations[42]. Together with ACC, it serves the neural processing of interpersonal motives[58].

Here, patients diagnosed with NPD and a group of healthy control subjects (CTL) were scanned using functional magnetic resonance imaging (fMRI) during a prosocial decision-making task. On each trial, participants had two options to split a monetary endowment between themselves and an unknown recipient. The splits systematically varied to manipulate the level of conflict between selfish and prosocial motives. Additionally, some trials allowed punishing the recipient by taking money from them. Concluding from previous studies in non-clinical samples with narcissistic traits [27–29], we expected lower generosity in the group of patients diagnosed with NPD than in CTL group, reflecting their decision making to be eventually governed by selfish motives. Additionally, we expected activity in neural conflict monitoring systems (i.e. dmPFC, ACC, SMC)[39] and differential RT patterns[34,36] to track the motivational conflict implied in the offer, potentially allowing to differentiate patients with NPD from CTL subjects.

## Methods

### Sample

A total of N=42 participants were included in the study. Participants belonging to the NPD group were recruited from Charité - Universitätsmedizin Berlin, Campus Benjamin Franklin, and were both inpatients and outpatients, but were included in the study only after being discharged from treatment. CTL group participants were recruited by announcements in the press and via publicly placed advertisements. The study was approved by the local ethics committee at Charité Berlin (EA4/092/10). Participants gave written informed consent and received monetary compensation for participating in the study. Structured Clinical Interviews for DSM IV axes I and II Disorders (SCID-I; SCID-II; 60,61; German Versions: 62,64) were administered to all participants by a trained clinical psychologist to validate NPD diagnosis and assess comorbidity (Supplementary Table 1). Potential participants for the CTL group were excluded if completion of SCID-I or SCID-II indicated the presence of any mental disorder. As assessed in prior studies of our group, interrater reliability (*κ*=0.80) and internal consistency (Cronbach’s *α*=0.86; sum of criteria) of SCID-II NPD diagnoses was acceptable [63]. Five participants were excluded from the analysis due to various reasons (Supplementary Methods), leaving a total of 37 participants (CTL: n=19, NPD: n=18) in the final analysis sample.

### Procedure

Before the study, a set of paper-pencil questionnaires were sent to all participants (Supplementary Table 1). Upon arrival at the MRI facility, all participants were briefed about the study protocol and gave written informed consent. They were informed that inside the MRI they were going to repeatedly decide how to share an endowment of 1000 points (equivalent to 10 €) between themselves and another person (henceforth: recipient) by choosing between two alternative splits of this endowment. To ensure interpersonal relevance of the task, they were informed that after task completion, a number of their choices would be selected randomly, defining how much money would be paid to them and to the next study participant (i.e. the recipient). Additionally, participants received money based on the choices of one other participant who was assigned randomly. All participants were tested alone and never met any other participant.

### Experimental Task

Stimulus presentation was performed using the *Presentation* software package (Neurobehavioral Systems, Albany, CA). On each trial, one option was displayed on the left and the other on the right side of the screen, with the amounts for the participant and the recipient on the top and the bottom, respectively (Figure 1). Participants chose by pressing a button with either the index or the middle finger. In a baseline (BL) condition, both options assigned precisely 500 points to each person. In five other conditions the level of conflict between selfish and prosocial motives was manipulated by systematically splitting the endowment, with one option favoring the participant more strongly (selfish option) than the other option (prosocial option). In one condition, options were very similar, inducing low motivational conflict (low conflict, LC). On other trials, one option assigned a high payoff to the participant, leaving little to the recipient, whereas the other option proposed a fairer split (medium conflict, MC). Still other trials did not allow to resolve motivational conflict this way, because avoiding selfishness required to choose an extremely prosocial option (high conflict, HC). Further, one condition allowed participants to increase their gain by subtracting money from the recipient, or to resort to a fairer split (medium conflict punishment, MCP). Finally, one condition also allowed participants to increase their payoff at the recipient’s cost, but the alternative would be to choose an extremely prosocial option (high conflict punishment, HCP). On each trial, following 500 ms fixation, a choice had to be made within the 3000 ms display of the options, otherwise the trial was coded as invalid. Each trial onset was triggered by an MRI pulse, and the inter-trial-interval (ITI) was jittered between 500 and 6500 ms in steps of 2000 ms (further information in Supplementary Methods).

**Figure 1.**
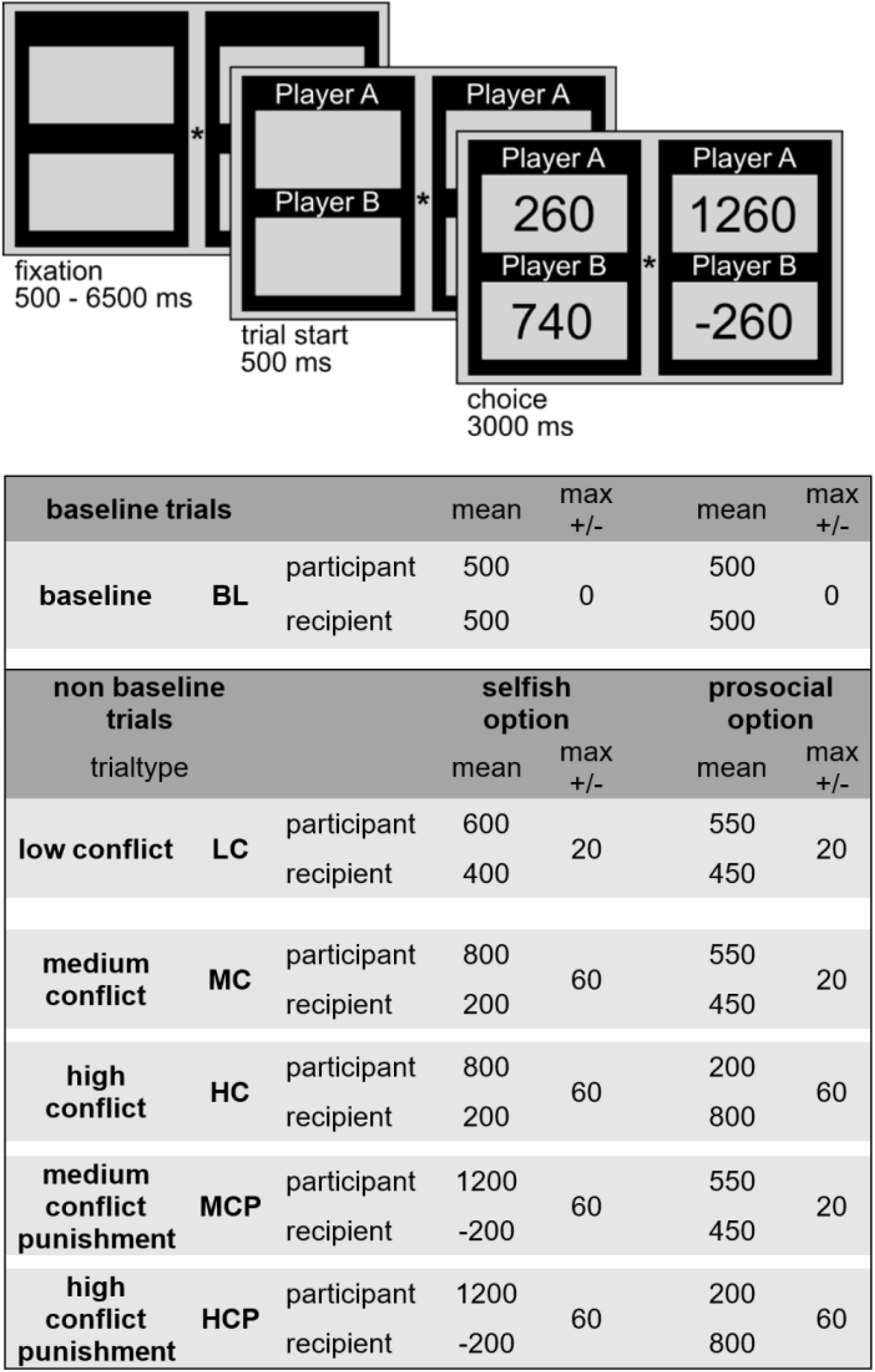
Structure of the experimental task. **Top**. Structure of a single trial. Following a jittered fixation (500-6500 ms), a trial was initialized for 500 ms and followed by a 3000 ms display of the two possible splits of the endowment of 1000 points (equivalent to 10€). Participants chose between options with a button press. Responses slower than 3000 ms were discarded. Player A is the participant and Player B the recipient. **Bottom**. Overview of all experimental conditions, with mean number of points assigned to each player and the preset maximum deviation from the mean assignment (max +/-). The split assigning most money to the participant was defined to be the selfish options, whereas the prosocial option was the one splitting the endowment in a relatively more beneficial way for the recipient. Note that due to this definition, the prosocial option did not necessarily assign more money to the recipient than to the participant. There were 12 different, pseudorandomized trial sequences that were designed to optimize BOLD efficiency. Thus, the number of trials in each sequence varied between 223 and 229 (see text for details).

### Statistical analyses of behavioral data

Statistical analyses of behavioral data were performed using JASP[64], jamovi[65], and the p.adjust() function in R[66]. We analyzed percentages of prosocial choices and median RTs for the different experimental conditions using repeated measures analysis of variance (ANOVA) and nonparametric tests, as appropriate. Since both choice options in the BL condition offered the same amount of 500 points to the player and the recipient, no selfish or prosocial option was defined and BL choice data were not analyzed. However, RTs from the BL condition were analyzed and it served as an explicit baseline for the analysis of fMRI data. To test the effects of increasing motivational conflict on behavior and neural activity, we compared HC trials against the average of LC and MC trials (LC&MC), because the latter both allowed to resolve the conflict with the same pro-social option in which the participant still received more payoff but amounts were rather similar (i.e. 550 and 450, respectively). This was not the case in the HC condition, in which the pro-social option implied a relative loss for the participant and gain for the recipient (200 and 800, respectively). Last, to test the main effect of possible punishment, we averaged MCP and HCP trials (PUN) and compared this to the average of MC and HC trials (noPUN).

### FMRI data acquisition and preprocessing

Echo-planar images (EPIs) were acquired on a Magnetom TrioTim Syngo (Siemens, Munich, Germany). The entire run consisted of 600 EPIs (37 ascending slices; 20% gap; 3*3*3 mm voxels; TR=2000 ms; TE=30 ms; FA=70°; FoV=192; GRAPPA=2). Additionally, a high-resolution T1-weighted MPRAGE image (176 ascending slices; 50% gap; 1*1*1 mm voxels; TR=1900 ms; TE=2.52 ms; FA=9°; FoV=256; GRAPPA=2) was acquired to improve spatial normalization. All fMRI data were preprocessed and analyzed using SPM 12[67] in Matlab R2019b (The Mathworks, Natick, Massachusetts, USA). EPIs were corrected for acquisition delay (slice timing), with the middle slice as reference, and for head motion (realignment). The MPRAGE image was coregistered to the mean EPI obtained from realignment and segmented using the unified segmentation algorithm implemented in SPM. EPIs were normalized to a common stereotactic reference frame (MNI) space using the transformation parameters obtained from segmentation, and smoothed using an 8 mm full-width-at-half-maximum isotropic Gaussian kernel.

### Statistical analysis of fMRI data

All fMRI data were analyzed using SPM12[67], using a two-stage mixed effects model. The first-level general linear model (GLM) contained six regressors of interest, one for the BL condition and one for each of the Choice conditions, plus the six rigid-body movement parameters obtained from realignment as regressors of no interest, to account for variance related to head motion, and a single constant. At the second level, we constructed a full-factorial ANOVA model, which included the contrast images of each Choice condition against the BL condition (e.g. HC>BL), separated by group, resulting in the within-subjects factor *condition* with five levels, and the between-subjects factor *group* (CTL, NPD).

To test for within-subject effects of RTs and prosocial choices, respectively, we set up two additional GLMs for each participant. Within each GLM, we modelled BL trials separately from Choice trials, and parametrically modulated, in this order, the latter with dummy predictors that indicated whether medium (MC, MCP) or high (HC, HCP) conflict trials were presented, and whether or not a given trial was a PUN trial (i.e., MCP, HCP). Last, we included either a predictor indicating the participant’s choice (prosocial=1; selfish=-1), or the RT on each trial (log-transformed and z-scored). Parametric modulators were orthogonalized. Additional regressors modelled invalid trials and head motion. One participant in the CTL group and three participants in the NPD group never chose the prosocial option and were therefore excluded from these analyses.

### Region-of-Interest Analysis

Since our primary focus was to investigate how motivational conflict engages regions associated with conflict monitoring, we used Neurosynth[68] to perform an automated meta-analysis of functional neuroimaging studies by searching for the term “conflict” and obtaining the association test map thresholded at *p*<.01 (false-discovery-rate-corrected; 337 studies). This map was then limited to include only clusters of 5 or more contiguous voxels and used as a region-of-interest, largely covering dorsal ACC, SMC, and aspects of bilateral AI (henceforth: *conflict mask*).

## Results

### Participants in NPD Group Acted More Selfishly, but Avoided Punishing Recipients

Across all Choice conditions, the NPD group (*MD*=9.80%, IQR=24.13%, *min*=0.0%, *max*=97.81%) chose the prosocial option less often than the CTL group (*MD*=45.20%, IQR=34.74%, *min*=0.0%, *max*=86.41%; Mann-Whitney-U test: *W*=260.5, one-sided *p*=.003, rank-biserial correlation=0.52, 90% *CI*=[0.26;0.72]; Figure 2 & Table 1).

**Table 1.** Behavioral data by group and experimental condition

| Choice behavior | CTL |  |  | NPD |  |  | W | one-sided *p*-value<sup>1</sup> | Rank-Biserial Correlation | 90% *CI* for Rank-Biserial Correlation |  |
| --- | --- | --- | --- | --- | --- | --- | --- | --- | --- | --- | --- |
| Condition | median % prosocial | [25<sup>th</sup>, 75<sup>th</sup> percentile] |  | median % prosocial | [25<sup>th</sup>, 75<sup>th</sup> percentile] |  |  |  |  | Lower | Upper |
| Overall | 45.20 | 35.30 | 70.03 | 9.80 | 1.38 | 25.51 | 260.5 | .003 | 0.52 | 0.26 | 0.72 |
| LC | 53.49 | 17.58 | 79.18 | 4.65 | 0 | 28.33 | 246 | .046 | 0.44 | 0.15 | 0.66 |
| MC | 53.33 | 29.35 | 88.04 | 2.30 | 0 | 19.99 | 273.5 | .005 | 0.60 | 0.36 | 0.77 |
| HC | 15.22 | 4.35 | 30.00 | 0.00 | 0 | 22.23 | 229 | .046 | 0.34 | 0.04 | 0.59 |
| MCP | 95.65 | 45.65 | 100.00 | 18.22 | 0 | 85.87 | 242 | .046 | 0.42 | 0.13 | 0.64 |
| HCP | 79.17 | 37.47 | 97.83 | 7.03 | 0 | 79.19 | 245 | .046 | 0.43 | 0.15 | 0.65 |

| Reaction times | CTL |  | NPD |  | *t* | *df* | two-sided *p*-value<sup>1</sup> | *d* | 95% *CI* for *d* |  |
| --- | --- | --- | --- | --- | --- | --- | --- | --- | --- | --- |
| Condition | *Mean* (ms) | *SD* | *Mean* (ms) | *SD* |  |  |  |  | Lower | Upper |
| Overall | 1138.63 | 231.72 | 951.50 | 218.88 | 2.52 | 35 | .016 | 0.83 | 0.15 | 1.50 |
| BL | 889.08 | 166.62 | 887.89 | 187.96 | 0.02 | 35 | .984 | 0.01 | -0.64 | 0.65 |
| LC | 1180.98 | 262.65 | 1060.97 | 283.77 | 1.34 | 35 | .571 | 0.44 | -0.22 | 1.09 |
| MC | 1164.63 | 301.72 | 950.72 | 275.30 | 2.25 | 35 | .124 | 0.74 | 0.07 | 1.40 |
| HC | 1310.61 | 362.42 | 976.31 | 258.75 | 3.21 | 35 | .017 | 1.06 | 0.36 | 1.74 |
| MCP | 1096.95 | 234.73 | 993.14 | 359.65 | 1.05 | 35 | .606 | 0.34 | -0.31 | 1.00 |
| HCP | 1393.84 | 414.36 | 1007.19 | 374.47 | 2.97 | 35 | .027 | 0.98 | 0.29 | 1.66 |

**Note.** CTL=Control group. NPD=Group diagnosed with Narcissistic Personality Disorder. CI=confidence interval. LC=Low Conflict. MC=Medium Conflict. HC=High Conflict. MCP=Medium Conflict Punishment. HCP=High Conflict Punishment.
<sup>1</sup>*p*-values for individual conditions (i.e. not "Overall") are Holm-corrected.

**Figure 2.**
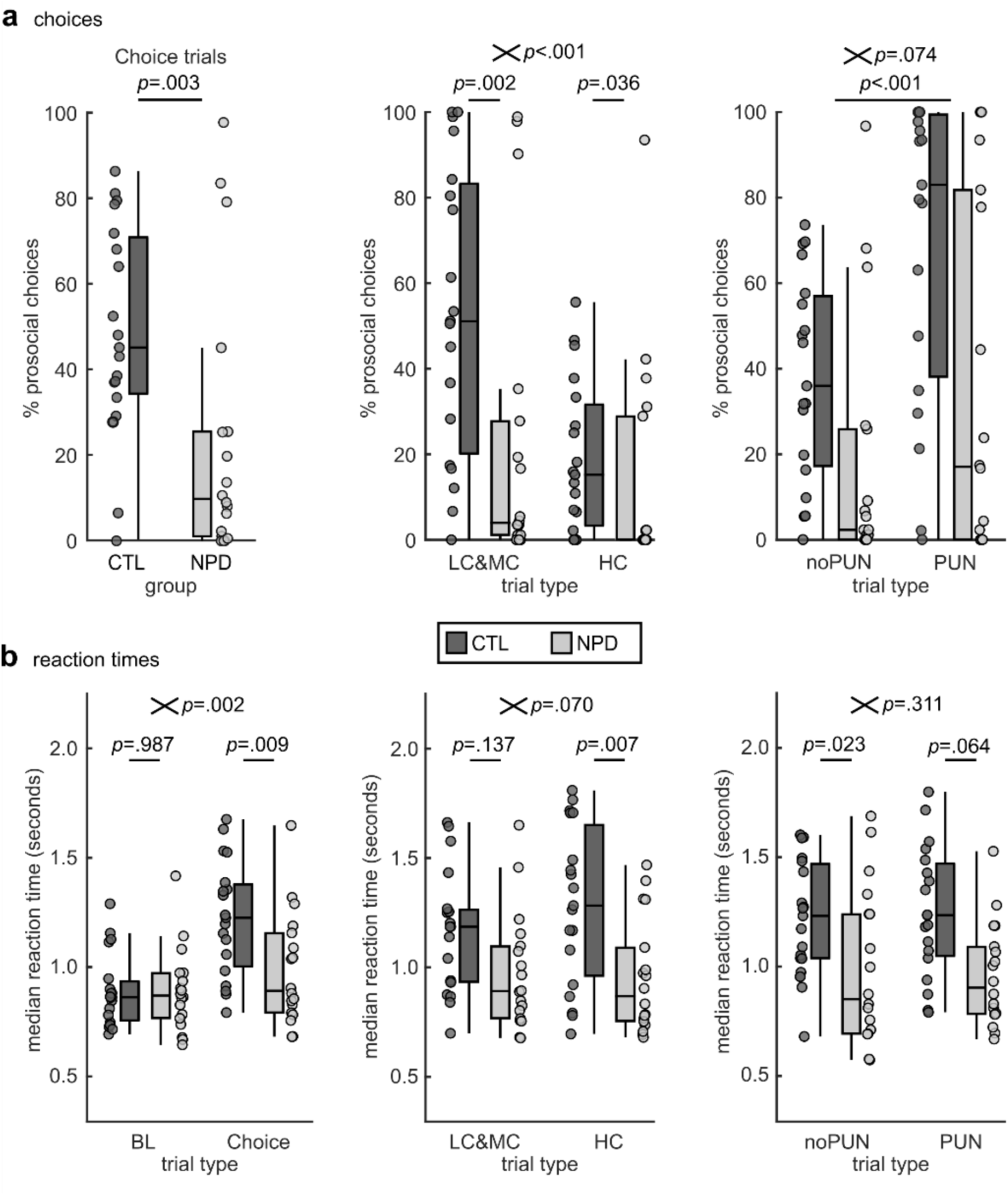
Behavioral data. **a**. Percentages of prosocial choices for the CTL group and NPD group overall (left), for trials with low and medium (LC&MC) vs high motivational conflict (HC; middle), and separately for trials without (noPUN: MC, HC), and with punishment (PUN: MCP, HCP). **b**. Median reaction times in seconds for BL and Choice trials (left), for LC&MC trials and HC trials (middle), and separately for PUN and noPUN trials. Where shown, small crossed bars represent interaction between group (CTL, NPD) and factor on x-axis. Central bars of boxplots show median, lower and upper box borders show 25^th^ and 75^th^ percentile and whiskers end at last data point within 1.5 times the interquartile range from lower or upper box border.

When motivational conflict was low, the CTL group behaved more prosocially than the NPD group (LC&MC, CTL group: *MD*=51.11%, *IQR*=59.53; NPD group: *MD*=3.97%, *IQR*=24.58; *W*=266.5, one-sided *p*=0.002, rank-biserial correlation=0.56, 90%, CI=[0.30;0.74]). When conflict increased, prosocial choices diminished in both groups (difference: LC&MC minus HC; CTL group: *MD*=29.69%, *IQR*=46.40, *W*=171.00, *p*<.001; NPD group: *MD*=1.20%, *IQR*=5.43, *W*=100.5, *p*=.003), and this effect was larger in the CTL group (*W*=288.50, two-sided *p*<.001, rank-biserial correlation=0.69, 95% *CI*=[0.43;0.84]). However, under HC, the CTL group still behaved more prosocially than the NPD group (CTL: *MD*=15.22%, *IQR*=25.65%; NPD: *MD*=0.00%, *IQR*=22.23%; *W*=229, one-sided, Holm-corrected *p*=0.046, rank-biserial correlation=0.34, 90% *CI*=[0.04;0.59]). Thus, the difference in prosociality between the groups was reduced, but still present, when prosocial behavior was very disadvantageous for the participant.

Across groups, prosociality increased when selfishness required to punish the recipient (difference PUN minus noPUN, CTL: *MD*=26.37%, *IQR*=48.80, *W*=158, one-sided *p*<.001, rank-biserial correlation=0.85, 90% *CI*=[0.67;0.93]; NPD: *MD*=1.63%, *IQR*=32.15, *W*=87, one-sided *p*=.016, rank-biserial correlation=0.66, 95% *CI*=[0.28;0.86]). This increase was descriptively, but not significantly, larger in the CTL than in the NPD group (*W*=219, one-sided *p*=.074, rank-biserial correlation=0.28, 90% *CI*=[-0.03;0.54]). Hence, having to punish others for one’s own benefit led to more prosocial behavior.

### Responses in NPD Group were Fast, but Slowed Down for Prosocial Decisions

RTs did not generally differ between groups (ANOVA with factors trial type [BL, Choice] and group [CTL, NPD]: *F*(1,35)=3.39, *p*=.074, 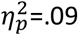). However, there was a significant main effect of trial type (*F*(1,35)=37.15, *p*<.001, 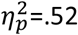), that was modulated by group (trial type*group interaction: *F*(1,35)=11.09, *p*=.002, 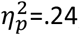; Figure 2). Post-hoc tests showed that the CTL group responded slower during Choice trials (*t*(54.2)=3.21, two-sided *p*_Holm_=.009), but there was no difference during BL trials (*t*(54.2)=0.02, two-sided *p*_Holm_=.987). Moreover, motivational conflict slowed RTs in the CTL group (Choice>BL: *t*(35)=6.76, *p*_Holm_<.001), but not in the NPD group (*t*(35)=1.93, *p*_Holm_=.186). Thus, both groups responsed comparably fast unless interpersonal motives needed to be reconciled, which slowed responding only in the CTL group.

Median RTs did not differ between selfish and prosocial decisions across groups (ANOVA with factors decision [selfish, prosocial] and group [CTL, NPD]: no main effect of decision: *F*(1,31)=4.03, *p*=.054, 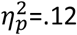; factor group: *F*(1,31)=2.91, *p*=.098, 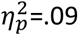). However, the effect of decision differed between groups (decision*group: *F*(1,32)=8.56, *p*=.006, 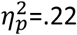; Figure 3), showing that the CTL group was slower than the NPD group during selfish choices (CTL>NPD: *t*(49.2)=2.93, *p_Holm_*=.026; Figure 3), but not during prosocial choices (CTL>NPD: *t*(49.2)=0.03, *p_Holm_*=1.000). Moreover, RTs within the CTL group did not differ between decisions (*t*(31)=0.68, *p_Holm_*=1.000), but the NPD group was faster for selfish than for prosocial decisions (*t*(31)=-3.34, *p_Holm_*=.013), which was not due to group-specific effects of the experimental conditions (Supplementary Table S4).

**Figure 3.**
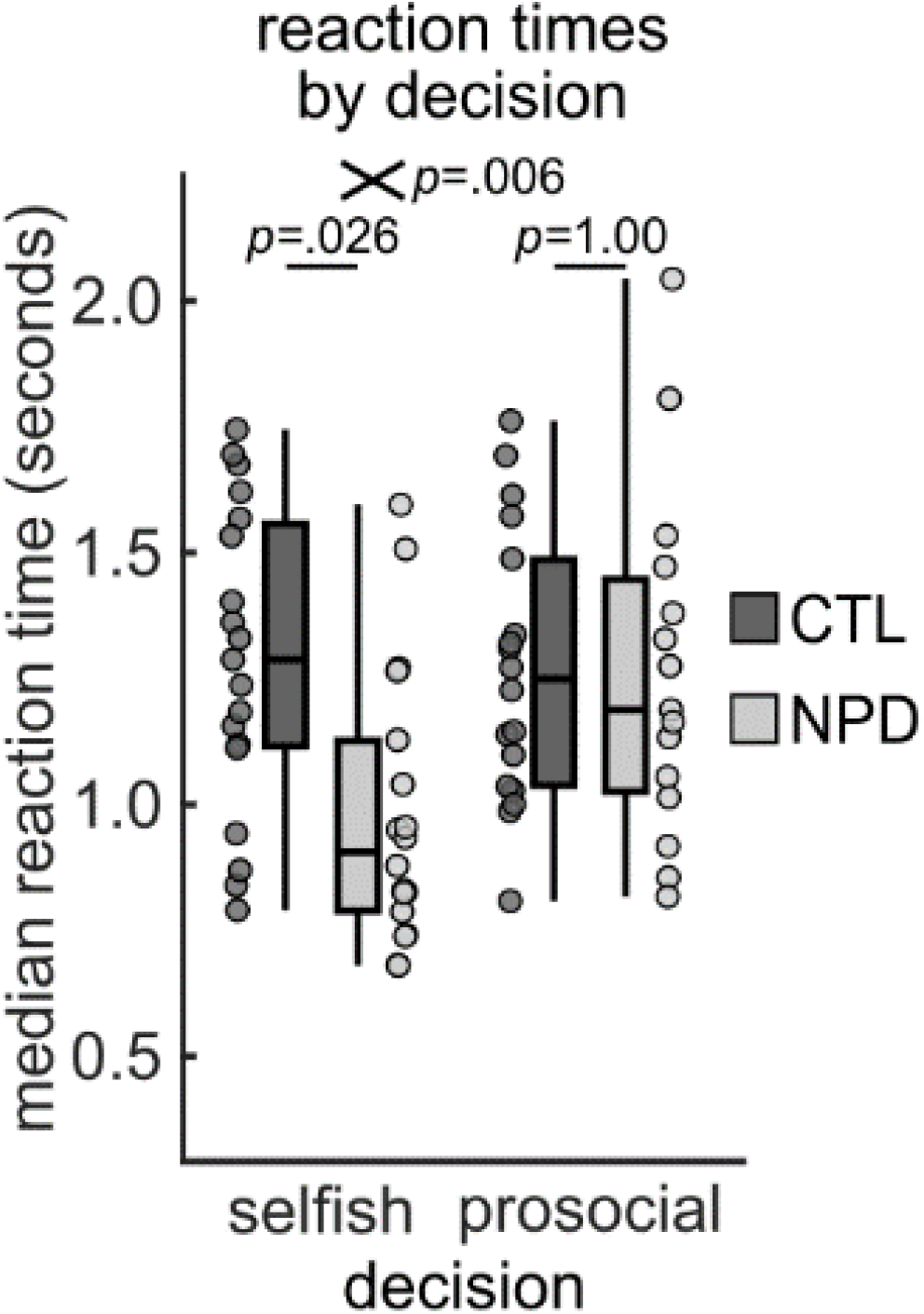
Reaction times separated by decision. Small crossed bar represents interaction between group (CTL, NPD) and decision made (selfish, prosocial). Small crossed bars represents interaction between group (CTL, NPD) and choice (selfish, prosocial). Central bars of boxplots show median, lower and upper box borders show 25^th^ and 75^th^ percentile and whiskers end at last data point within 1.5 times the interquartile range from lower or upper box border.

RTs were insensitive to level of motivational conflict across groups (ANOVA with factors level of conflict [LC&MC, HC] and group [CTL, NPD]: *F*(1,35)=1.86, *p*=.181, 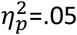). Yet, we found a main effect of group (*F*(1,35)=8.53, *p*=.006, 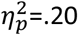), that was driven by the HC condition (CTL>NPD: *t*(35)=3.21, *p*_Holm_=.017). The interaction of both factors was not significant (*F*(1,35)=3.50, *p*=.070, 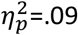). Thus, when prosociality was disadvantageous for the participants, responses in the CTL group were slower than in the NPD group.

Last, the option to punish did not impact reaction times across groups (ANOVA with factors punishment [noPUN, PUN] and group [CTL, NPD]: *F*(1,35)=0.00, *p*=.983, 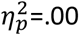). Again, the CTL group responded slower than the NPD group (main effect of group: *F*(1,35)=8.30, *p*=.007, 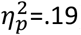), which was due to noPUN trials (CTL>NPD: *t*(43.2)=3.06, *p*_Holm_=.023). No significant interaction was found (punishment*group: *F*(1,35)=1.06, *p*=.311, 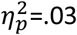).

### Conflict-Related Neural Activity During Prosocial Decision Making Was Reduced in NPD Group

Across groups, presence of motivational conflict (Choice>BL) elicited activity in right dorsal midcingulate cortex (MNI x,y,z: 6,30,42; *T*=5.79, *k*=68, *p*_FWE_=.001; whole-brain), right dorsolateral prefrontal cortex (dlPFC; 46,34,22; *T*=6.45, *k*=146, *p*_FWE_<.001), bilateral parietal cortex (right: 28,-60,44; *T*=9.32, *k*=1833, *p*_FWE_<.001; left: -26,-70,32; *T*=9.63, *k*=1349, *p*_FWE_<.001), and further regions (see Supplementary Information for full activation coordinates). This effect was stronger for the CTL than the NPD group in left dorsomedial prefrontal cortex (dmPFC; -6,30,38; *T*=3.86, *k*=29, *p*_FWE_=.023) and left preSMA (−4,12,52; *T*=4.44, *k*=10, *p*_FWE_=.003; corrected inside the *conflict* mask, Figure 4), while the opposite comparison (i.e. Choice>BL * NPD>CTL) did not yield any significant results, indicating that considering potential interpersonal consequences is linked to conflict-related neural activity particularly in the CTL group.

**Figure 4.**
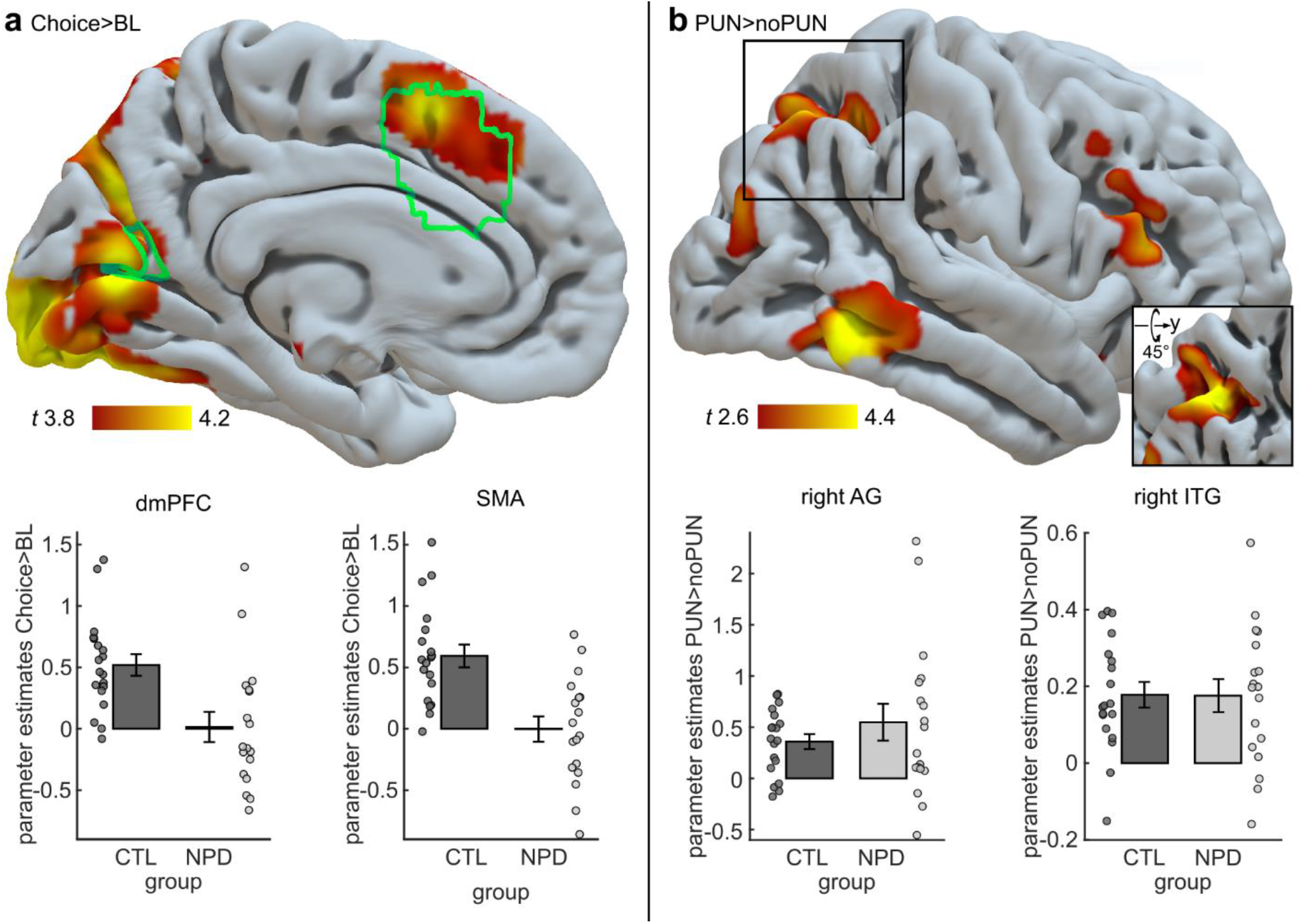
Neural activity during the experimental task. **a**. Neural activity in regions associated with the term *conflict* (green outline) including dorsomedial prefrontal cortex (dmPFC) and supplementary motor area (SMA) was higher for the contrast Choice>BL (*p*<.05, whole-brain FWE-corrected; displayed at *p*<.0001, uncorrected). This effect was larger in the CTL group than in the NPD group, visualized in the bar plots. Neural activation data in bar plots are based on mean parameter estimate across all voxels in dmPFC and SMA, respectively, in which the interaction of Choice>BL * CTL>NPD was significant at *p*<.05, small-volume FWE-corrected inside the *conflict* mask). **b**. Neural activity in right angular gyrus (AG) and right inferior temporal gyrus (ITG) was higher for PUN than for noPUN trials, but did not differ between groups. Bar plots show mean parameter estimates across all voxels in right AG and right ITG in which the contrast PUN>noPUN was significant at *p*<.05, whole-brain FWE-corrected (displayed at *p*<.005, uncorrected). Inset in bottom right corner depicts activation in right AG, rotated around the y axis by 45 degrees. Errorbars are +/- 1 standard error of the mean.

Contrasting HC against the average of LC and MC (HC>LC&MC) to test for effects of increasing motivational conflict on neural activity did not yield any significant effects or interactions.

When selfish behavior required punishing others (PUN>noPUN), activity increased in right AG (34,-68,52; *T*=5.35, *k*=73, *p*_FWE_=.006; whole-brain), right inferior temporal gyrus (ITG; 54,-56,-10; *T*=5.24, *k*=19, *p*_FWE_=.009), and right superior parietal cortex (26,-72,54; *T*=4.84, *k*=1, *p*_FWE_=.043). The effect in right ITG was significant when performing this analysis in the CTL group alone (48,-56,-8; *T*=5.20, *k*=33, *p*_FWE_=.004; whole-brain), which also showed significantly stronger activations for PUN vs noPUN trials in left AI (−32,18,-14; *T*=5.25, *k*=14, *p*_FWE_=.009), right inferior frontal gyrus (50,20,16; *T*=4.89, *k*=3, *p*_FWE_=.037), and right middle temporal gyrus (62,-44,2; *T*=5.26, *k*=35, *p*_FWE_=.008). However, despite no significant effects in the NPD group, PUN>noPUN did not differ between groups. Hence, when selfish behavior was possible only by punishing others, this induced stronger activity in brain regions like right AG and ITG, which was driven by the CTL group.

### Opportunities to Punish Increased Conflict-Related Neural Activity Across Groups

To predict task-related neural activity with individual differences in prosociality, we regressed the first-level Choice>BL contrast images on the participants’ mean percentages of prosocial choices while controlling for group differences. Across groups, higher prosociality predicted increasing activity in right dlPFC (36,38,14; *T*=5.80, *k*=3, *p*_FWE_=.040; whole-brain), but no group differences were found. Regressing the contrast HC>LC&MC on the corresponding difference in prosociality (i.e., HC minus LC&MC) did not yield any significant effects. However, increasing prosociality from noPUN to PUN trials predicted corresponding increases in neural activity in right SMA (6,6,48; *T*=6.16, *k*=3, *p*_FWE_=.018; whole-brain) across groups. Thus, higher overall prosociality predicted neural activity in right dlPFC, and increased prosociality in order not to punish others was linked to increased SMA activity across groups.

Since participants in the NPD group showed low overall prosociality and responded more slowly during prosocial than during selfish choices, we expected stronger trial-by-trial conflict-related neural activity for prosocial choices in the NPD than in the CTL group[34,36,39], but no such effect was found.

### Reaction Times Predicted Conflict-Related Neural Activity Within and Across Participants

Last, under motivational conflict, slower participants showed higher activity in right mid cingulate gyrus (Choice>BL: 4,32,36; *T*=5.37, *k*=53, *p*_FWE_=.001; corrected inside the *conflict* mask), left AI (−38,18,2; *T*=5.87, *k*=22, *p*_FWE_<.001), and left SMA (−4,14,50; *T*=5.37, *k*=111, *p*_FWE_=.001). Similarly, slower RTs due to increasing conflict were associated with higher neural activity in SMA (HC>LC&MC: 2,16,44; *T*=6.56, *k*=224, *p*_FWE_<.001; corrected inside the *conflict* mask), and left (−32,16,2; *T*=5.9, *k*=16, *p*_FWE_<.001) and right AI (40,16,0; *T*=4.73, *k*=6, *p*_FWE_=.008). Additionally, slower trial-by-trial RTs predicted neural activity in ACC, bilateral AI, and lateral prefrontal and parietal cortices, and an exploratory *F*-test indicated that trial-by-trial RTs were less predictive of mid cingulate neural activity in the NPD than in the CTL group (−6,26,36; *F*=18.05, *k*=1, *p*_FWE_=.048; corrected inside the *conflict* mask).

## Discussion

In the present study we investigated how prosocial decision making and underlying neural processes in patients diagnosed with NPD are shaped by conflicting interpersonal motives. Compared to control subjects, the NPD group behaved less prosocially, responded faster, and showed less neural activity in regions associated with conflict monitoring such as the SMA and cingulate cortex[38,39]. When prosociality was disadvantageous for the participants, it diminished in both groups, although control subjects still behaved more prosocially than the NPD group. Both groups were similarly affected by the requirement to punish others for one’s own benefit, which shifted behavior away from selfishness, and increased neural activity in right AG and ITG. Last, when participants in the NPD group acted prosocially, thus deviating from their selfish mode of behavior, this slowed responding to a level comparable to the CTL group.

Our finding of low generosity in participants diagnosed with NPD is well in line with existing literature from non-clinical narcissistic samples[27,28]. We hypothesized that this may either result from more readily available tendencies towards selfishness[35] or, alternatively, from cognitively effortful arbitration between conflicting interpersonal motives; i.e. either to selfishly increase one’s own gain or to behave prosocially in order to be seen in a good light by others[9,10]. Our findings speak to the former hypothesis, as the mainly selfish responses in the NPD group were generally fast, which was paralleled by comparably low conflict-related neural activity[38,39], and rare prosocial choices were considerably slower. Participants in the CTL group more readily utilized the fairness information provided in each trial to guide their decisions, causing motivational conflict and the need to engage in cognitive control[34,39]. In sum, our data nicely coincide with earlier research showing that deviations from selfish or prosocial habits are marked by slowing RTs[36]. Thus, using response times one may reveal individual differences in social value orientations[35], with participants in the NPD group being more strongly oriented toward selfishness.

Notably, behavior in the NPD group was not fully insensitive to its potential social consequences. Similar to the CTL group, participants in the NPD group acted more selfishly when prosociality was believed to be highly detrimental, and acted more prosocially when their own benefit was conditional on punishing others. Prosocial behavior can be shaped by inducing prosocial motives[58], and explicit instructions to consider others’ perspectives can increase prosocial responding in non-clinical narcissistic individuals[69]. Although we did not provide such explicit instructions, our task was effective for pitting selfish against prosocial motives. Notably, recipients were anonymous, which is known to impact behavior in prosocial decision games[70], and did not have an option to reciprocate, invalidating any potential economic repercussions of unfair behavior. Despite these circumstances, patients diagnosed with NPD showed some responsiveness to others’ potential needs, underlining that pathological narcissism does not equate to an absolute incapacity, but perhaps a reduced propensity to act prosocially[71]. This finding points to the potential of empathy interventions to increase perspective taking[72] for increasing prosocial behavior[58], which could be reflected in neural activity in networks mediating interpersonal motivations[32,58,71]. Such interventions could entail practicing to actively consider how one’s social behavior affects others, and to imagine how others will respond during interpersonal interactions. In consequence, such therapeutic interventions could elicit higher prosociality and more favorable evaluations from others, stabilizing self-esteem[46,73] and overall well-being[74].

Our findings also speak to neural underpinnings of prosociality more generally. Precisely, neural activity in SMA, which is linked to cognitive control of behavior[38], was higher for participants who favored prosociality over punishing others for their own benefit. When such punishment was possible, this increased activity in AG, which may reflect more specifically social-cognitive processes like moral reasoning[55], or domain-general cognitive processes like attentional reorienting[75] that may also be engaged during social behavior. In addition, ITG was activated when punishment was possible, which has previously been associated with fairness considerations[42]. Last, participants with higher overall levels of prosociality showed increased activity in dlPFC, which was also responsive to the general presence of conflicting interpersonal motives, alongside other regions of the cognitive control network (i.e. dmPFC, AI, ACC, SMA, and parietal cortex).

So far, few studies have investigated the neural correlates of interpersonal behavior in NPD using experimental paradigms in combination with fMRI. By doing so, our data contribute to a more detailed understanding of NPD in terms of impairments in interpersonal functioning, as introduced by DSM-5, section 3 [1]. Collectively, our data indicate that selfish behavior in NPD relies on diminished consideration of prosocial motives, which obviates the need to resolve conflict with selfish tendencies. However, this may reflect a reduced propensity, not ability, to act prosocially[69,71]. Potentially, this reflects differences between NPD and, for instance, antisocial personality disorder, which could be tested explicitly in future studies. In terms of treatment implications, our results point to possibilities for enhancing prosocial behavior and thereby improving social relationships and well-being in NPD. Regarding social cognition and interpersonal behavior more broadly, our findings support the notion that these functions rely on domain-general control processes [38–40], that draw on contextual social information to decide between generous or selfish behavior[43].

## Data Availability

The data of this study are available from the corresponding author upon request.

## Acknowledgements

We would like to thank Sylvia Richter and Thomas Münte for helpful discussions during development of the task and Michael Scholz for his help with the optimization of the trial sequence for the fMRI task. We further thank Jürgen Baudewig, Ricardo Heydenblut, and Christian Kainz for assistance with MRI data acquisition. Research leading to this publication has been funded by the Deutsche Forschungsgemeinschaft (Sören Krach; KR 3803/9-1; Aline Vater and Stefan Roepke; EXC 302; Björn Schott; SFB 779, TP A08).

## Supplementary Information

Supplementary information is available at MP’s website.

