## Supplementary Information for "Reduced Frontal Cortical Tracking of Conflict between Selfish versus Prosocial Motives in Narcissistic Personality Disorder"

### Supplementary Methods

#### Sample

Five subjects were excluded from the analyses: because the MRI scan was interrupted prematurely (n=3); due to technical problems no log file was recorded (n=1); participant never responded in one condition (n=1).

#### Experimental task

In the first of the Choice conditions, the low conflict (LC) condition, the selfish option assigned an average of 600 points to the participant and 400 points to the recipient, while the prosocial option assigned an average of 550 points to the participant and 450 points to the recipient. The values on each trial randomly deviated up to 20 points from the average amount. That is, if the participant decided selfishly this would not entail a strong detriment for the recipient. In the medium conflict (MC) condition, the prosocial option was the same as in LC, but the selfish option split the endowment more unfairly (participant: 800 points, recipient: 200 points, up to 60 points deviation). In the high conflict (HC) condition, the selfish option assigned 800 points to the participant and 200 to the recipient, with a deviation of up to 60 points, and the prosocial option provided the opposite split. That is, deciding selfishly would be very beneficial for the participant, but very unfair to the recipient, and vice versa. In the fifth condition (medium conflict + punishment, MCP), participants could choose whether they wanted to receive 550 points and give 450 to the recipient (with up to 20 points deviation), or whether they would rather take 1200 points and subtract 200 points from the recipient (with up to 60 points deviation), thereby punishing the other. Here, participants could decide very selfishly with a strong detriment for the recipient, or evade this option in favor of a relatively fair split. In the sixth condition, (high conflict + punishment; HCP), participants again could choose between a selfish option equivalent to the one in the MCP condition (participant: 1200; recipient: -200), but the alternative was to receive only 200 points and give 800 to the recipient (with up to 60 points deviation from the average amount). That is, deciding not to punish the recipient would come at a high monetary cost for the participant, inducing high conflict between prosocial and selfish motives.

The experimental paradigm was optimized by estimating the statistical efficiency of different ITIs and jitters(67), thereby creating twelve different trial sequences, with a maximum of three consecutive repetitions of a single trial type. Each sequence was used similarly often in both groups ($X^2(11)=3.51$, $p=.982$; Supplementary Table S2). This optimization procedure caused slight variability between sequences in the number of trials per condition and the total number of trials (Supplementary Table S3), but these did not differ between groups (all *p*s>.290), and neither did the number of valid trials (all *p*s>.406).

### Supplementary Tables

**Supplementary Table S1.** Demographics and psychometric characteristics of study samples

| | CTL group (n=19) | | NPD group (n=18) | | | | |
|---|---|---|---|---|---|---|---|
| | *M* | *SD* | *M* | *SD* | *t* | *p (2-sided)* | |
| age | 38.84 | 10.5 | 38.94 | 2.14 | -.03 | .975 | |
| | count | | count | | *X²* | *p* (2-sided) | |
| female/male | 5/14 | | 5/13 | | 0.01 | .920 | |
| **Axis I diagnosis** | 0 | | 9 | | | | |
| Depressive episode (mild) | 0 | | 2 | | | | |
| Depressive episode (moderate) | 0 | | 3 | | | | |
| Recurrent depressive disorder (moderate) | 0 | | 1 | | | | |
| Recurrent depressive disorder (currently in remission) | 0 | | 3 | | | | |
| **Axis II diagnosis** | | | | | | | |
| NPD | 0 | | 18 | | | | |
| BPD | 0 | | 6 | | | | |
| APD | 0 | | 1 | | | | |
| HPD | 0 | | 1 | | | | |
| | *M* | *SD* | *M* | *SD* | *t* | *p (2-sided)* | |
| **PNI total** | 2.37 | 0.81 | 3.97 | 0.70 | -6.47 | <.001 | |
| Contingent self-esteem | 2.34 | 1.12 | 4.07 | 0.94 | -5.10 | <.001 | |
| Exploitative | 2.21 | 1.03 | 4.02 | 1.19 | -4.95 | <.001 | |
| Self-sacrificing Self-enhancement | 2.78 | 1.25 | 3.64 | 1.22 | -2.11 | .042 | |
| Hiding the Self | 2.71 | 1.08 | 4.37 | 0.89 | -5.06 | <.001 | |
| Grandiose Fantasy | 2.15 | 0.83 | 3.83 | 1.05 | -5.42 | <.001 | |
| Devaluing | 2.11 | 0.73 | 3.83 | 1.06 | -5.78 | <.001 | |
| Entitlement Rage | 2.30 | 0.93 | 3.94 | 1.06 | -5.05 | <.001 | |
| **NPI total** | 8.95 | 3.31 | 23.00 | 6.60 | -8.12 | <.001 | a |
| Leadership | 2.05 | 1.22 | 7.28 | 2.52 | -7.96 | <.001 | a |
| Grandiosity/exhibitionism | 2.00 | 1.63 | 4.39 | 1.91 | -4.09 | <.001 | |
| Entitlement/exploitative | 1.00 | 0.88 | 2.83 | 1.10 | -5.51 | <.001 | |
| **BDI** | 2.95 | 4.62 | 20.78 | 9.93 | -6.94 | <.001 | a |

**Note**. CTL=control group. NPD=Narcissistic Personality Disorder. BPD=Borderline Personality Disorder. APD=Anxious Personality Disorder. HPD=Histrionic Personality Disorder.
a=Welch test was performed instead of student's t-test due to unequal variances.

**Supplementary Table S2.** Number of task versions used in each group

| | group | | |
|---|---|---|---|
| **trial sequence version** | **CTL** | **NPD** | **Total** |
| 1 | 2 | 2 | 4 |
| 2 | 2 | 2 | 4 |
| 3 | 2 | 3 | 5 |
| 4 | 1 | 0 | 1 |
| 5 | 2 | 1 | 3 |
| 6 | 1 | 2 | 3 |
| 7 | 2 | 1 | 3 |
| 8 | 2 | 1 | 3 |
| 9 | 2 | 1 | 3 |
| 10 | 1 | 2 | 3 |
| 11 | 1 | 1 | 2 |
| 12 | 1 | 2 | 3 |
| Total | 19 | 18 | 37 |

| ***N*** | ***X²*** | ***df*** | ***p*** |
|---|---|---|---|
| 37 | 3.51 | 11 | .982 |

**Supplementary Table S3.** Number of trials of each condition, separately for different trial sequence versions of the experimental task

| trial sequence version | BL | LC | MC | HC | MCP | HCP |
|---|---|---|---|---|---|---|
| **1** | 43 | 45 | 45 | 45 | 23 | 23 |
| **2** | 44 | 46 | 46 | 46 | 24 | 23 |
| **3** | 45 | 45 | 45 | 44 | 22 | 22 |
| **4** | 44 | 45 | 45 | 45 | 23 | 23 |
| **5** | 44 | 46 | 46 | 46 | 23 | 24 |
| **6** | 44 | 46 | 46 | 46 | 23 | 23 |
| **7** | 45 | 45 | 45 | 45 | 23 | 23 |
| **8** | 45 | 46 | 46 | 46 | 23 | 23 |
| **9** | 44 | 46 | 46 | 46 | 23 | 24 |
| **10** | 44 | 46 | 46 | 46 | 23 | 23 |
| **11** | 44 | 46 | 46 | 46 | 24 | 23 |
| **12** | 45 | 45 | 45 | 44 | 22 | 22 |

**Supplementary Table S4.** Hierarchical linear model predicting reaction times (in log-transformed milliseconds)

| predictor | *b* | *SE* | *df* | *t* | *p* | |
|---|---|---|---|---|---|---|
| Constant (CTL, LC, selfish choice) | 7.05 | 0.05 | 41.44 | 144.07 | <.001 | *** |
| isNPD | -0.15 | 0.07 | 40.98 | -2.13 | .039 | * |
| isMC | -0.02 | 0.02 | 6621.21 | -1.51 | .132 | |
| isHC | 0.09 | 0.02 | 6625.39 | 5.46 | <.001 | *** |
| isMCP | -0.07 | 0.02 | 6622.74 | -3.74 | <.001 | *** |
| isHCP | 0.09 | 0.02 | 6621.89 | 4.62 | <.001 | *** |
| choice was prosocial | 0.04 | 0.01 | 6657.28 | 2.75 | .006 | ** |
| isNPD * isMC | -0.07 | 0.02 | 6621.23 | -3.10 | .002 | ** |
| isNPD * isHC | -0.13 | 0.02 | 6624.88 | -5.66 | <.001 | *** |
| isNPD * isMCP | -0.04 | 0.03 | 6624.40 | -1.62 | .105 | |
| isNPD * isHCP | -0.15 | 0.03 | 6622.19 | -5.75 | <.001 | *** |
| isNPD * choice was prosocial | 0.12 | 0.02 | 6616.20 | 5.23 | <.001 | *** |

**Note.** Fixed effects predictors were group, condition, choice, and the two two-way interactions between group and each of the other two factors. Model included random intercepts for each subject.

### Full activation coordinates

**Supplementary Table S5.** CHOICE>BL across groups, whole-brain FWE-corrected at p<.05

| set | | cluster | | | | peak | | | | | mm | | |
|---|---|---|---|---|---|---|---|---|---|---|---|---|---|
| p | c | p(FWE) | p(FDR) | k | p(unc) | p(FWE) | p(FDR) | T | Z | p(unc) | x | y | z |
| <.001 | 19 | <.001 | <.001 | 1735 | <.001 | <.001 | <.001 | 14.48 | 65535.00 | <.001 | 32 | -90 | -6 |
| | | <.001 | <.001 | 1984 | <.001 | <.001 | <.001 | 13.45 | 65535.00 | <.001 | -28 | -90 | -8 |
| | | | | | | <.001 | <.001 | 8.74 | 65535.00 | <.001 | -44 | -76 | -12 |
| | | | | | | <.001 | <.001 | 8.58 | 7.82 | <.001 | -44 | -66 | -14 |
| | | <.001 | <.001 | 1349 | <.001 | <.001 | <.001 | 9.63 | 65535.00 | <.001 | -26 | -70 | 32 |
| | | | | | | <.001 | <.001 | 8.41 | 7.69 | <.001 | -26 | -62 | 50 |
| | | <.001 | <.001 | 1833 | <.001 | <.001 | <.001 | 9.32 | 65535.00 | <.001 | 28 | -60 | 44 |
| | | | | | | <.001 | <.001 | 9.10 | 65535.00 | <.001 | 32 | -70 | 30 |
| | | | | | | <.001 | <.001 | 8.39 | 7.68 | <.001 | 22 | -70 | 56 |
| | | <.001 | <.001 | 629 | <.001 | <.001 | <.001 | 8.28 | 7.59 | <.001 | -4 | -74 | -26 |
| | | | | | | <.001 | <.001 | 8.01 | 7.38 | <.001 | 6 | -74 | -26 |
| | | | | | | <.001 | <.001 | 7.29 | 6.80 | <.001 | -6 | -76 | -38 |
| | | <.001 | <.001 | 146 | <.001 | <.001 | .001 | 6.45 | 6.10 | <.001 | 46 | 34 | 22 |
| | | | | | | <.001 | .238 | 5.21 | 5.01 | <.001 | 48 | 44 | 18 |
| | | <.001 | <.001 | 161 | <.001 | <.001 | .002 | 6.41 | 6.07 | <.001 | -40 | 2 | 30 |
| | | <.001 | .003 | 76 | .002 | <.001 | .002 | 6.38 | 6.04 | <.001 | 0 | -56 | -36 |
| | | <.001 | .014 | 49 | .008 | <.001 | .005 | 6.16 | 5.85 | <.001 | -6 | 14 | 54 |
| | | <.001 | .000 | 178 | .000 | <.001 | .012 | 5.95 | 5.67 | <.001 | -2 | -74 | 8 |
| | | .011 | .269 | 9 | .213 | <.001 | .014 | 5.92 | 5.64 | <.001 | -22 | -38 | -40 |
| | | <.001 | .005 | 68 | .003 | <.001 | .024 | 5.79 | 5.53 | <.001 | 6 | 30 | 42 |
| | | | | | | .004 | .100 | 5.44 | 5.22 | <.001 | 6 | 22 | 44 |
| | | .001 | .034 | 35 | .022 | .001 | .036 | 5.68 | 5.44 | <.001 | 44 | -36 | 48 |
| | | .004 | .114 | 19 | .078 | .006 | .145 | 5.34 | 5.14 | <.001 | -28 | -64 | -30 |
| | | .005 | .140 | 16 | .103 | .007 | .164 | 5.31 | 5.10 | <.001 | -32 | -68 | -42 |
| | | .029 | .633 | 2 | .567 | .026 | .574 | 4.98 | 4.81 | <.001 | 56 | 16 | 36 |
| | | .018 | .417 | 5 | .351 | .026 | .574 | 4.97 | 4.80 | <.001 | 44 | 46 | -12 |
| | | .035 | .699 | 1 | .699 | .027 | .575 | 4.96 | 4.79 | <.001 | 22 | -38 | -40 |
| | | .035 | .699 | 1 | .699 | .047 | .936 | 4.82 | 4.66 | <.001 | 22 | 40 | -16 |

**Supplementary Table S6.** CHOICE>BL * CTL>NPD, FWE-corrected at p<.05 inside the conflict mask

| set | | cluster | | | | peak | | | | | mm | | |
|---|---|---|---|---|---|---|---|---|---|---|---|---|---|
| p | c | p(FWE) | p(FDR) | k | p(unc) | p(FWE) | p(FDR) | T | Z | p(unc) | x | y | z |
| .001 | 2 | .008 | .295 | 29 | .147 | .003 | .280 | 4.44 | 4.31 | <.001 | -4 | 12 | 52 |
| | | | | | | .007 | .364 | 4.18 | 4.08 | <.001 | 2 | 12 | 52 |
| | | .020 | .390 | 10 | .390 | .023 | .670 | 3.86 | 3.78 | <.001 | -6 | 30 | 38 |
| | | | | | | .027 | .670 | 3.81 | 3.73 | <.001 | -2 | 30 | 40 |
| | | | | | | .045 | .898 | 3.66 | 3.59 | <.001 | 0 | 26 | 42 |

**Supplementary Table S7.** PUN>noPUN across groups, whole-brain FWE-corrected at p<.05

| set | | cluster | | | | peak | | | | | mm | | |
|---|---|---|---|---|---|---|---|---|---|---|---|---|---|
| p | c | p(FWE) | p(FDR) | k | p(unc) | p(FWE) | p(FDR) | T | Z | p(unc) | x | y | z |
| <.001 | 3 | <.001 | .006 | 73 | .002 | .006 | .264 | 5.35 | 5.14 | <.001 | 34 | -68 | 52 |
| | | .004 | .117 | 19 | .078 | .009 | .264 | 5.24 | 5.04 | <.001 | 54 | -56 | -10 |
| | | .035 | .699 | 1 | .699 | .043 | .853 | 4.84 | 4.69 | <.001 | 26 | -72 | 54 |

**Supplementary Table S8.** PUN>noPUN in CTL group, whole-brain FWE-corrected at p<.05

| set | | cluster | | | | peak | | | | | mm | | |
|---|---|---|---|---|---|---|---|---|---|---|---|---|---|
| p | c | p(FWE) | p(FDR) | k | p(unc) | p(FWE) | p(FDR) | T | Z | p(unc) | x | y | z |
| <.001 | 4 | .001 | .050 | 33 | .025 | .004 | .223 | 5.41 | 5.20 | <.001 | 48 | -56 | -8 |
| | | .001 | .050 | 35 | .022 | .008 | .223 | 5.26 | 5.06 | <.001 | 62 | -44 | 2 |
| | | .006 | .167 | 14 | .125 | .009 | .223 | 5.25 | 5.05 | <.001 | -32 | 18 | -14 |
| | | .024 | .475 | 3 | .475 | .037 | .726 | 4.89 | 4.72 | <.001 | 50 | 20 | 16 |

**Supplementary Table S9.** First-level contrast CHOICE>BL regressed on the overall percentage of prosocial choices, whole-brain FWE-corrected at p<.05

| set | | cluster | | | | peak | | | | | mm | | |
|---|---|---|---|---|---|---|---|---|---|---|---|---|---|
| p | c | p(FWE) | p(FDR) | k | p(unc) | p(FWE) | p(FDR) | T | Z | p(unc) | x | y | z |
| .050 | 1 | .019 | .375 | 3 | .375 | .040 | .803 | 5.80 | 4.78 | <.001 | 36 | 38 | 14 |

**Supplementary Table S10.** First-level contrast PUN>noPUN regressed on corresponding difference in % prosocial choices, whole-brain FWE-corrected at p<.05

| set | | cluster | | | | peak | | | | | mm | | |
|---|---|---|---|---|---|---|---|---|---|---|---|---|---|
| p | c | p(FWE) | p(FDR) | k | p(unc) | p(FWE) | p(FDR) | T | Z | p(unc) | x | y | z |
| .050 | 1 | .018 | .361 | 3 | .361 | .018 | .348 | 6.16 | 4.99 | <.001 | 6 | 6 | 48 |

**Supplementary Table S11.** First-level contrast CHOICE>BL regressed on corresponding difference in median reaction times, FWE-corrected at p<.05 inside the conflict mask

| set | | cluster | | | | peak | | | | | mm | | |
|---|---|---|---|---|---|---|---|---|---|---|---|---|---|
| p | c | p(FWE) | p(FDR) | k | p(unc) | p(FWE) | p(FDR) | T | Z | p(unc) | x | y | z |
| <.001 | 4 | .008 | .209 | 22 | .157 | <.001 | .073 | 5.87 | 4.82 | <.001 | -38 | 18 | 2 |
| | | <.001 | .017 | 111 | .004 | .001 | .081 | 5.37 | 4.52 | <.001 | -4 | 14 | 50 |
| | | | | | | .002 | .088 | 5.19 | 4.40 | <.001 | 6 | 18 | 48 |
| | | | | | | .003 | .088 | 5.10 | 4.35 | <.001 | 4 | 16 | 52 |
| | | .002 | .072 | 53 | .036 | .001 | .081 | 5.37 | 4.52 | <.001 | 4 | 32 | 36 |
| | | | | | | .002 | .081 | 5.31 | 4.48 | <.001 | 6 | 36 | 34 |
| | | | | | | .003 | .088 | 5.07 | 4.33 | <.001 | 0 | 30 | 40 |
| | | | | | | .011 | .275 | 4.59 | 4.00 | <.001 | -2 | 30 | 36 |
| | | | | | | .023 | .511 | 4.30 | 3.81 | <.001 | 4 | 28 | 42 |
| | | .040 | .790 | 1 | .790 | .046 | .918 | 4.03 | 3.61 | <.001 | -24 | 20 | 4 |

**Supplementary Table S12.** First-level contrast HC>LC&MC regressed on corresponding difference in median reaction times, FWE-corrected at p<.05 inside the conflict mask

| set | | cluster | | | | peak | | | | | mm | | |
|---|---|---|---|---|---|---|---|---|---|---|---|---|---|
| p | c | p(FWE) | p(FDR) | k | p(unc) | p(FWE) | p(FDR) | T | Z | p(unc) | x | y | z |
| <.001 | 4 | <.001 | <.001 | 224 | <.001 | <.001 | .006 | 6.56 | 5.21 | <.001 | 2 | 16 | 44 |
| | | | | | | .006 | .184 | 4.86 | 4.19 | <.001 | 8 | 24 | 30 |
| | | | | | | .009 | .210 | 4.70 | 4.09 | <.001 | -6 | 22 | 42 |
| | | | | | | .016 | .306 | 4.47 | 3.93 | <.001 | -4 | 24 | 38 |
| | | .010 | .410 | 16 | .205 | .000 | .024 | 5.90 | 4.84 | <.001 | -32 | 16 | 2 |
| | | | | | | .002 | .083 | 5.23 | 4.43 | <.001 | -32 | 24 | 4 |
| | | .022 | .439 | 6 | .439 | .001 | .048 | 5.53 | 4.62 | <.001 | -26 | 22 | 2 |
| | | .022 | .439 | 6 | .439 | .008 | .210 | 4.73 | 4.10 | <.001 | 40 | 16 | 0 |

**Supplementary Table S13.** Main effect of trial-by-trial reaction times (log-transformed and z-scored), whole-brain FWE-corrected at p<.05

| set | | cluster | | | | peak | | | | | mm | | |
|---|---|---|---|---|---|---|---|---|---|---|---|---|---|
| p | c | p(FWE) | p(FDR) | k | p(unc) | p(FWE) | p(FDR) | T | Z | p(unc) | x | y | z |
| <.001 | 27 | <.001 | <.001 | 10371 | <.001 | <.001 | <.001 | 13.29 | 65535.00 | <.001 | -30 | -2 | 60 |
| | | | | | | <.001 | <.001 | 11.79 | 7.44 | <.001 | -28 | -54 | 48 |
| | | | | | | <.001 | <.001 | 11.72 | 7.42 | <.001 | -48 | -32 | 50 |
| | | <.001 | <.001 | 2308 | <.001 | <.001 | <.001 | 10.79 | 7.12 | <.001 | -6 | 12 | 50 |
| | | | | | | <.001 | <.001 | 10.75 | 7.10 | <.001 | 4 | 24 | 42 |
| | | | | | | <.001 | <.001 | 10.37 | 6.97 | <.001 | -8 | 16 | 42 |
| | | <.001 | <.001 | 722 | <.001 | <.001 | <.001 | 10.61 | 7.05 | <.001 | -6 | -74 | -26 |
| | | | | | | <.001 | <.001 | 10.30 | 6.94 | <.001 | 4 | -76 | -20 |
| | | | | | | <.001 | <.001 | 10.23 | 6.92 | <.001 | -6 | -76 | -34 |
| | | <.001 | <.001 | 2282 | <.001 | <.001 | <.001 | 9.98 | 6.82 | <.001 | 32 | 22 | 0 |
| | | | | | | <.001 | <.001 | 9.77 | 6.74 | <.001 | 42 | 34 | 24 |
| | | | | | | <.001 | .002 | 8.54 | 6.24 | <.001 | 32 | 12 | 2 |
| | | <.001 | <.001 | 683 | <.001 | <.001 | .000 | 9.94 | 6.81 | <.001 | -30 | 22 | -2 |
| | | | | | | <.001 | .001 | 8.99 | 6.43 | <.001 | -32 | 18 | -12 |
| | | | | | | <.001 | .009 | 7.79 | 5.89 | <.001 | -32 | 16 | 8 |
| | | <.001 | <.001 | 114 | <.001 | <.001 | .001 | 8.64 | 6.28 | <.001 | 2 | -56 | -34 |
| | | <.001 | <.001 | 277 | <.001 | <.001 | .002 | 8.61 | 6.27 | <.001 | -34 | -58 | -32 |
| | | <.001 | <.001 | 837 | <.001 | <.001 | .002 | 8.42 | 6.19 | <.001 | 40 | -64 | -16 |
| | | | | | | <.001 | .004 | 8.12 | 6.05 | <.001 | 44 | -78 | -10 |
| | | | | | | <.001 | .005 | 8.03 | 6.01 | <.001 | 54 | -56 | -10 |
| | | <.001 | <.001 | 602 | <.001 | <.001 | .009 | 7.78 | 5.89 | <.001 | 4 | -66 | 8 |
| | | | | | | <.001 | .018 | 7.41 | 5.71 | <.001 | -12 | -74 | 6 |
| | | | | | | <.001 | .035 | 7.10 | 5.55 | <.001 | 14 | -76 | 16 |
| | | <.001 | <.001 | 339 | <.001 | <.001 | .016 | 7.46 | 5.73 | <.001 | -38 | -70 | -16 |
| | | | | | | <.001 | .029 | 7.17 | 5.59 | <.001 | -44 | -62 | -8 |
| | | | | | | <.001 | .367 | 6.09 | 4.99 | <.001 | -40 | -76 | -2 |
| | | <.001 | <.001 | 372 | <.001 | <.001 | .019 | 7.38 | 5.69 | <.001 | -40 | 54 | 10 |
| | | | | | | <.001 | .089 | 6.69 | 5.33 | <.001 | -34 | 58 | 4 |
| | | | | | | <.001 | .147 | 6.49 | 5.22 | <.001 | -44 | 46 | -4 |
| | | <.001 | <.001 | 102 | <.001 | <.001 | .021 | 7.33 | 5.67 | <.001 | 24 | -54 | -20 |
| | | <.001 | <.001 | 196 | <.001 | <.001 | .023 | 7.30 | 5.65 | <.001 | 8 | -24 | -8 |
| | | | | | | <.001 | .023 | 7.28 | 5.64 | <.001 | -4 | -24 | -16 |
| | | | | | | .002 | .050 | 6.93 | 5.46 | <.001 | 10 | -14 | -8 |
| | | <.001 | .008 | 39 | .004 | .001 | .039 | 7.04 | 5.52 | <.001 | -8 | -14 | -2 |
| | | .001 | .037 | 22 | .025 | .002 | .049 | 6.95 | 5.47 | <.001 | -6 | -28 | -4 |
| | | .005 | .138 | 11 | .097 | .006 | .154 | 6.47 | 5.21 | <.001 | 8 | -36 | -28 |
| | | .011 | .271 | 6 | .211 | .009 | .206 | 6.34 | 5.14 | <.001 | 30 | 46 | -12 |
| | | <.001 | .021 | 29 | .012 | .011 | .245 | 6.27 | 5.10 | <.001 | -42 | 30 | 14 |
| | | .008 | .205 | 8 | .152 | .011 | .252 | 6.25 | 5.09 | <.001 | 36 | 28 | 42 |
| | | <.001 | .030 | 25 | .018 | .018 | .381 | 6.06 | 4.98 | <.001 | 28 | 56 | 6 |
| | | .001 | .035 | 23 | .022 | .022 | .472 | 5.98 | 4.93 | <.001 | 44 | 52 | -4 |
| | | .031 | .624 | 1 | .624 | .026 | .543 | 5.91 | 4.89 | <.001 | -22 | -60 | 0 |
| | | .016 | .374 | 4 | .305 | .031 | .640 | 5.84 | 4.85 | <.001 | 22 | -64 | 2 |
| | | .024 | .555 | 2 | .473 | .041 | .833 | 5.73 | 4.78 | <.001 | -36 | 36 | 6 |
| | | .031 | .624 | 1 | .624 | .045 | .919 | 5.69 | 4.75 | <.001 | -28 | -68 | -42 |
| | | .031 | .624 | 1 | .624 | .048 | .969 | 5.66 | 4.74 | <.001 | -14 | -72 | -6 |
| | | .031 | .624 | 1 | .624 | .049 | .977 | 5.66 | 4.74 | <.001 | -32 | -66 | -42 |

**Supplementary Table S14.** Test for group difference of trial-by-trial reaction times (log-transformed and z-scored) on neural activity, FWE-corrected at p<.05 inside the conflict mask

| set | | cluster | | | | peak | | | | | mm | | |
|---|---|---|---|---|---|---|---|---|---|---|---|---|---|
| p | c | p(FWE) | p(FDR) | k | p(unc) | p(FWE) | p(FDR) | F | Z | p(unc) | x | y | z |
| .050 | 1 | .039 | .768 | 1 | .768 | .048 | .962 | 18.05 | 3.61 | <.001 | -6 | 26 | 36 |

### Supplementary Figures

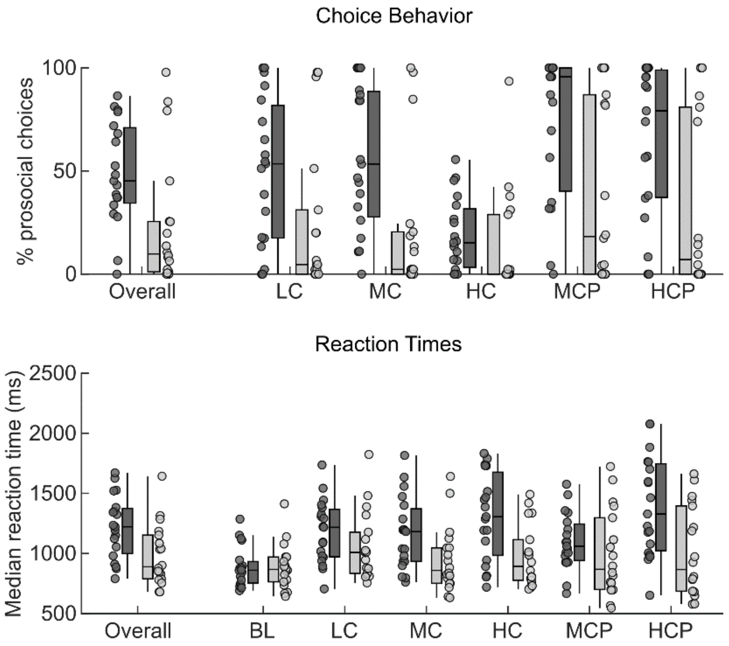


**Figure S1.** Choice behavior and response times. **Top.** Percentages of prosocial choices for the two groups overall (left), and separately for all conditions except BL, where prosocial and selfish decisions are undefined. **Bottom.** Median reaction times for the two groups overall (left) and separately for all conditions. BL=Baseline. LC=Low Conflict. MC=Medium Conflict. HC=High Conflict. MCP=Medium Conflict Punishment. HCP=High Conflict. Median reaction times in milliseconds across BL and Choice trials, separately for groups. Lines within boxes show the median, upper/lower box borders the 75th and 25th percentiles, and whisker length is 1.5 * interquartile range. Bright grey depicts data for NPD group, and dark grey for the CTL group.
